# Genetic profile and functional proteomics of anal squamous cell carcinoma: proposal for a molecular classification

**DOI:** 10.1101/19009522

**Authors:** Lucía Trilla-Fuertes, Ismael Ghanem, Angelo Gámez-Pozo, Joan Maurel, Laura G-Pastrián, Marta Mendiola, Cristina Peña, Rocío López-Vacas, Guillermo Prado-Vázquez, Elena López-Camacho, Andrea Zapater-Moros, Victoria Heredia, Miriam Cuatrecasas, Pilar García-Alfonso, Jaume Capdevila, Carles Conill, Rocío García-Carbonero, Ricardo Ramos-Ruiz, Claudia Fortes, Carlos Llorens, Paolo Nanni, Juan Ángel Fresno Vara, Jaime Feliu

## Abstract

**Background:** Anal squamous cell carcinoma is a rare tumor. Chemo-radiotherapy yields a 50% 3-year relapse-free survival rate in advanced anal cancer, so improved predictive markers and therapeutic options are needed.

**Methods:** High-throughput proteomics and whole-exome sequencing were performed in 46 paraffin samples from anal squamous cell carcinoma patients. Hierarchical clustering was used to establish groups *de novo*. Then, probabilistic graphical models were used to study the differences between groups of patients at the biological process level.

**Results:** A molecular classification into two groups of patients was established, one group with increased expression of proteins related to adhesion, T lymphocytes and glycolysis; and the other group with increased expression of proteins related to translation and ribosomes. The probabilistic graphical model showed that these two groups presented differences in metabolism, mitochondria, translation, splicing and adhesion processes. Additionally, these groups showed different frequencies of genetic variants in some genes, such as *ATM, SLFN11* and *DST*. Finally, genetic and proteomic characteristics of these groups suggested the use of some possible targeted therapies, such as PARP inhibitors or immunotherapy.

**Conclusions:** In this study, a molecular classification of anal squamous cell carcinoma using high-throughput proteomics and whole-exome sequencing data was proposed. Moreover, differences between the two established groups suggested some possible therapies.

## Background

Anal squamous cell carcinoma (ASCC) is a relatively rare cancer. In the United States there are 8,000 new estimated cases per year, accounting for approximately 2.7% of all gastrointestinal cancers. Of these, more than 1,000 cases will result in death(1).

Since the 1970s, the standard treatment has consisted of the combination of 5-fluorouracil (5FU), mitomycin C or cisplatin and radiotherapy (2). This treatment is particularly effective in T1/T2 tumors, achieving complete regression in 80-90% of cases. However, in advanced anal cancers (T3-4 or N+), the disease-free survival (DFS) rate is only around 50% (3). Therefore, due to the lack of advances in the last years, new therapeutic strategies are needed to improve these outcomes.

Whole-exome sequencing (WES) focused on the identification of disease-causing genes is now being implemented into clinical practice (4). The first work that announced entire exome sequencing was published by Ng et al (5). Since then, personalized medicine has focused on identifying the cause of rare diseases and cancers.

With the recent improvements in mass-spectrometry (MS) techniques, high-throughput proteomics has made it possible to identify thousands of proteins (6). Proteins are the effectors of biological processes, being closer to the phenotype than genes or transcripts. On the other hand, probabilistic graphical models (PGMs) were successfully used in previous studies to characterize tumors from a functional perspective (7-9). Moreover, when used in combination, proteomics and genomics provide complementary information.

Previous studies in ASCC were focused in the characterization of genetic variants in this disease using next-generation sequencing techniques. The most frequent mutated genes, such as *PIK3CA, FBXW7, FAT1* or *ATM*, were characterized (10-13). On the other hand, Herfs et al. used MS proteomics in microdissected anal samples to establish differential protein expression patterns depending of the location (squamous or transitional) (14). However, until date, a molecular classification of ASCC has not been established

In this study, we combined WES with high-throughput proteomics to further characterize a cohort of 46 ASCC tumors. This is the first time that a combined study of these characteristics in ASCC has been done. Genomics provides information about the genetic causes of disease and proteins are the ultimate effectors of biological processes. Therefore, a study of these two –omics allows us to obtain a broader picture of the molecular features of ASCC tumors.

## Methods

### Patient cohort

Forty-six formalin-fixed, paraffin-embedded (FFPE) samples from patients diagnosed with ASCC were analyzed by WES and MS proteomics. The study was approved by the Ethical Committee of Hospital Universitario La Paz. Informed consent was obtained for all patients in the study. Samples were reviewed by an experienced pathologist and all the samples included at least 70% invasive tumor cells. Patients were required to have a histologically-confirmed diagnosis of ASCC, be 18 years of age or older; have an Eastern Cooperative Oncology Group performance status (ECOG-PS) of 0 to 2; have received no prior radiotherapy or chemotherapy for this malignancy and present with no metastasis. Demographic information related to the tumor and the treatments was collected. Human papilloma virus (HPV) infection was determined by CLART® HPV2 (Genomica).

### DNA isolation

One 10 mm section from each FFPE sample was deparaffinized and DNA was extracted using GeneRead DNA FFPE Kit (Qiagen), in accordance with the manufacturer’s instructions. Once eluted, the DNA was frozen at −80 °C until use.

### Protein isolation

Proteins were extracted from FFPE samples as previously described (15). Briefly, FFPE sections were deparaffinized in xylene and washed twice with absolute ethanol. Protein extracts were prepared in a 2% SDS buffer by a protocol based on heat-induced antigen retrieval. Protein concentration was measured using the MicroBCA Protein Assay Kit (Pierce-Thermo Scientific). Protein extracts (10 µg) were digested with trypsin (1:50) and SDS was removed from the digested lysates using Detergent Removal Spin Columns (Pierce). Peptides were desalted using self-packed C18 stage tips, dried and resolubilized with 10 µl of 3% acetonitrile, 0.1% formic acid.

### Whole-exome sequencing experiments

WES from 46 ASCC FFPE samples was performed. The isolated DNA was evaluated by Picogreen and mean size was controlled by gel electrophoresis. Genomic DNA was divided by mechanical methods (Bioruptor) to a mean size of approximately 200 bp. At that point, DNA tests were fixed, phosphorylated, A-followed and ligated to explicit connectors, trailed by PCR-interceded naming with Illumina-explicit successions and test explicit standardized identifications (Kapa DNA library age unit).

Exome capture was performed utilizing the VCRome framework (catch size of 37 Mb, Nimblegen) under a multiplexing of 8 tests for every capture response. Capture was performed entirely in accordance to Nimblegen enhancement instructions. After the capture, libraries were cleansed, measured and titrated using Real Time PCR before sequencing. Tests were then sequenced to a surmised inclusion of 4.5 Gb per test in Illumina-NextSeq NS500 (Illumina Inc.) utilizing 150 cycles (2×75) High Output cartridges.

Raw data files were available in Sequence Read Archive (SRA, https://www.ncbi.nlm.nih.gov/sra) under the name PRJNA573670.

### Bioinformatics analyses of whole-exome sequencing data

The quality of the WES experiments was verified using FASTQC (http://www.bioinformmatics.babraham.ac.uk/projects/fastqc). First, primers were removed using Cutadapt. Then, FASTQ files were filtered by quality using PrinSeq. Both tools are available in GPRO tool (16). Sequence alignment was performed using the human genome h19 as the reference and BWA tools (17), Samtools (18) and Picard Tools (http://picard.sourceforge.net). Variant calling was performed using the MuTect tool from the GATK4 package (19) combined with PicardTools, first, to create a panel of normal samples (PON) and second, to perform the variant calling (20). The PON was built using 11 samples from Iberian exomes from a 1000genomes database (http://www.ncbi.nlm.nih.gov/sra/).

Finally, variants were annotated using Variant Effect Predictor (VEP) (21). The information about the genetic variants provided by VEP was used to filter the genetic variants. The filtering criteria were: a frequency in the general population, according gnomAD database, of less than 1%, a high or moderate impact, and the presence of a variant of this gene in our cohort in at least 10% of the patients.

### Liquid chromatography-mass spectrometry analysis

MS analysis was performed using a Q Exactive HF-X mass spectrometer (Thermo Scientific) equipped with a Digital PicoView source (New Objective) and coupled to a M-Class UPLC (Waters). Solvent composition at the two channels was 0.1% formic acid for channel A and 0.1% formic acid, 99.9% acetonitrile for channel B. For each sample 3 μL of peptides were loaded on a commercial MZ Symmetry C18 Trap Column (100Å, 5 µm, 180 µm × 20 mm, Waters) followed by nanoEase MZ C18 HSS T3 Column (100Å, 1.8 µm, 75 µm × 250 mm, Waters). The peptides were eluted at a flow rate of 300 nL/min by a gradient from 8 to 27% B in 85 min, 35% B in 5 min and 80% B in 1 min. Samples were acquired in a randomized order. The mass spectrometer was operated in data-dependent acquisition mode (DDA), acquiring full-scan MS spectra (350−1’400 m/z) at a resolution of 120’000 at 200 m/z after accumulation to a target value of 3’000’000, followed by HCD (higher-energy collision dissociation) fragmentation on the twenty most intense signals per cycle. HCD spectra were acquired at a resolution of 15’000 using normalized collision energy of 28 and a maximum injection time of 22 ms. The automatic gain control (AGC) was set to 100’000 ions. Charge state screening was enabled. Singly, unassigned, and charge states higher than seven were rejected. Only those precursors with an intensity above 110’000 were selected for MS/MS. Precursor masses previously selected for MS/MS measurement were excluded from further selection for 30 s, and the exclusion window was set at 10 ppm. The samples were acquired using internal lock mass (22) calibration on m/z 371.1012 and 445.1200.

The MS proteomics results were handled using the local laboratory information management system (LIMS) (22) and all relevant data have been deposited to Chorus under the project name “Anal squamous cell carcinoma proteomics”.

### Protein identification and label-free protein quantification

The acquired raw MS data was processed by MaxQuant (version 1.6.2.3), followed by protein identification using the integrated Andromeda search engine (23). Spectra were searched against a Uniprot reference proteome (taxonomy 9606, canonical version from 2016-12-09), concatenated to its reversed decoyed fasta database and common protein contaminants. Methionine oxidation and N-terminal protein acetylation were set as variable modifications. Enzyme specificity was set to trypsin/P allowing for a minimal peptide length of 7 amino acids and a maximum of two missed-cleavages. MaxQuant Orbitrap default search settings were used. The maximum false discovery rate (FDR) was set to 0.01 for peptides and 0.05 for proteins. Label-free quantification was enabled and a 2 minutes window for match between runs was applied. In the MaxQuant experimental design template, each file is kept separate in the experimental design to obtain individual quantitative values.

As quality criteria, the detectable measurement in at least 75% of the samples and the presence of two unique peptides were applied. Log2 of the data was calculated and missing values were imputed to a normal distribution using Perseus software (24).

### Probabilistic graphical models and functional node activity

With the aim of studying proteomics data from a functional point of view, PGMs compatible with high-dimensional data, were used. *grapHD* (25) and R v3.2.5 were used to generate the PGM from proteomics expression without any *a priori* information, based on correlation as associative measurement. The network was built in two steps: first, the spanning tree with maximum likelihood was found and, then, the edges was chosen based on the reduction of the Bayesian Information Criteria (BIC) and the preservation of the decomposability of the graph (26). The resulting network was analyzed to define a functional structure by gene ontology analyses, as in previous works (7-9). Briefly, the branches of the network were analyzed by gene ontology and a majority function for each branch was assigned, thereby determining different functional nodes in the network. Gene ontology analyses were performed using DAVID 6.8 webtool (27) using “homo sapiens” as background and GOTERM-FAT, Biocarta and KEGG as categories.

Once each branch had been assigned a function, functional node activities were calculated as the mean of the proteins of each branch related to the main function of that branch (8, 9). Then, comparisons between groups using Mann-Whitney test were done.

### Metabolic modeling and estimation of tumor growth rate

Flux Balance Analysis (FBA) is a method used to model the flow of metabolites through biochemical networks (28). It allows the growth rate or production rate of a given metabolite to be estimated using as input gene or protein expression data. In this study we used the whole human reconstruction Recon2 and the biomass reaction included in this model as the objective function and as representative of tumor growth (29). Proteomics data was introduced into the model to make accurate predictions by solving Gene-Protein-Reaction rules (GPRs), which contain the relationships between genes and enzymes, using a modified algorithm of Barker et al. (30) and a modified E-flux (7, 31). FBA calculations were performed using the COBRA Toolbox library, available for MATLAB (32).

### Statistical analyses

Statistical analyses were performed in GraphPad Prism 6 and SPSS IBM Statistics 20. Network analyses were performed using Cytoscape software. Hierarchical cluster and Significance Analysis of Microarrays (SAM) were performed using MeV software. SAM analysis allows the identification of differential proteins between groups by a t-test corrected by permutations over the number of samples. The significance was determined using the False Discovery Rate (FDR) (33). The Genomics of Drug Sensitivity in Cancer database (https://www.cancerrxgene.org/) was used to find possible therapeutic targets. P-values are two-sided and considered statistically significant under 0.05.

## Results

### Patient cohort

Forty-six patients diagnosed with non-metastatic ASCC were recruited for this study. Twenty-eight patients came from the VITAL clinical trial (GEMCAD-09-02, NCT01285778), treated with panitumumab, 5FU, Mitomycin C, and radiotherapy. The other 18 patients were included from the routine clinical practice at Hospital Universitario La Paz and Hospital Clinic and were treated with cisplatin-5FU or Mitomycin C-5FU, and concomitant radiotherapy.

For the survival analyses, 4 patients that could not receive chemo-radiotherapy were excluded (two of them had stage I anal carcinomas, and the other two had stage III tumors). The median follow-up was 33.18 months (5.53-116.4) and there are 13 relapse events. All clinical characteristics are shown in Table 1.

### Whole-exome sequencing experiments

Forty-six FFPE samples were analyzed by WES. The mean coverage of the samples was 42.6x, with the exception of one sample that presented a coverage of 3.57x. This sample was dismissed from the subsequent analyses. Once this sample was dismissed, all the samples presented a mapping efficiency of 90-98%, with the exception of one sample (with a mapping efficiency of 75.4%). Human exome has > 195,238 exonic regions, of which only 23,021 (11.21% of the human exome) have not been mapped in any sample.

After VEP analysis and filtering, 382 genes that presented a genetic variant with high or moderate impact in at least 10% of our cohort were identified (Sup Table 1). These genes were mostly related to DNA repair, chromatin binding and focal adhesion processes. *PIK3CA* was mutated in the 40% of the patients of our cohort, *FBXW7* in 16%, *FAT1* in 18%, and *ATM* was mutated in 27% of the patients. Figure 1 summarizes the high and moderate impact alteration landscape in ASCC.

**Figure 1:**
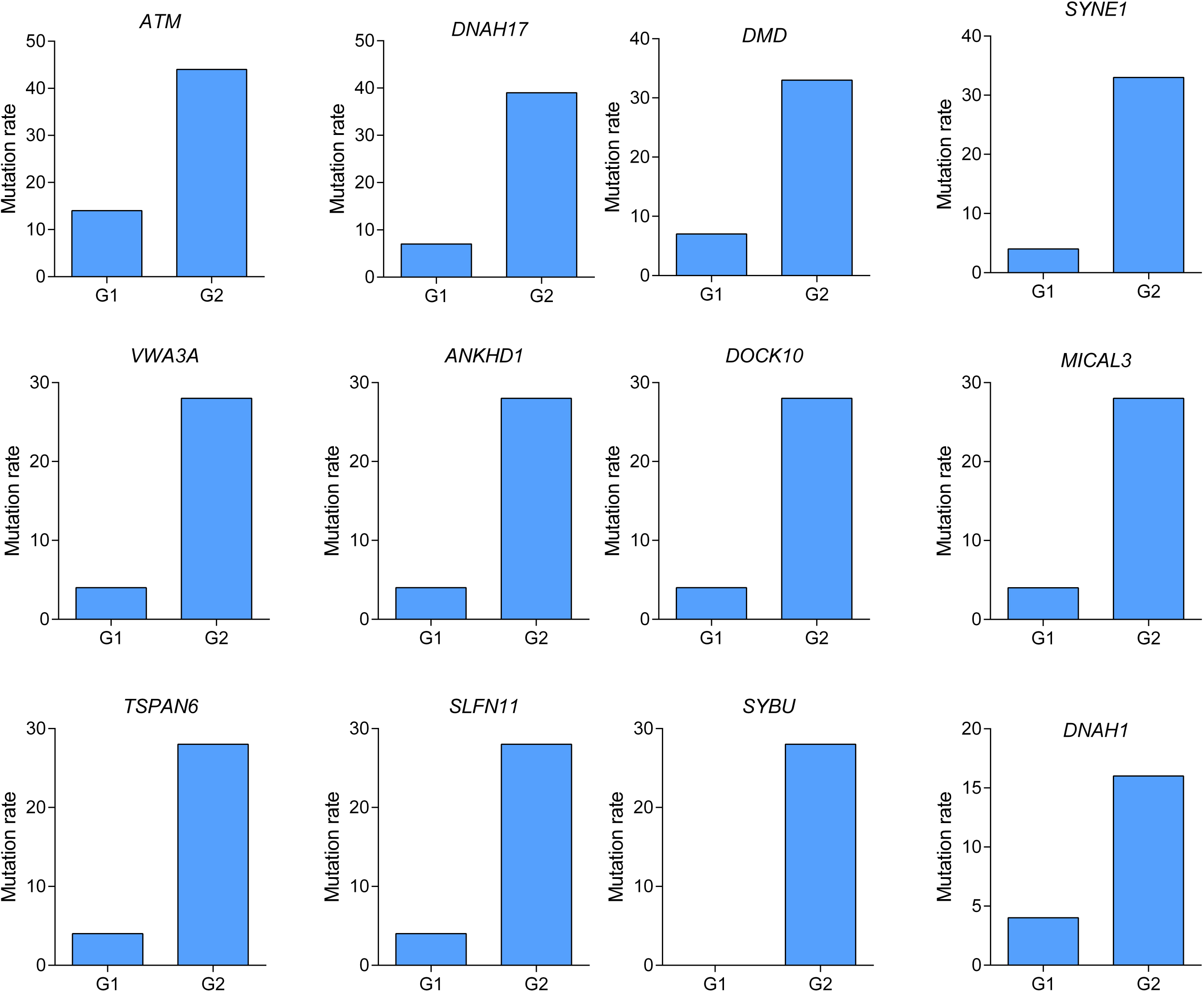
High and moderate impact genetic variants located in genes mutated in at least 20% of the ASCC patients of this cohort.

### Proteomics experiments

After dismissing one sample in the WES experiments, 45 FFPE samples were analyzed by MS and 6,035 proteins were identified. After applying quality criteria (detectable measurement in at least 75% of the samples and at least two unique peptides), 1,954 proteins were used for the subsequent analyses.

### De novo identification of groups based on differential proteomics profiles

With the aim of defining *de novo* molecular groups of patients, a hierarchical cluster was used. Two different molecular groups were obtained. A SAM was performed to define the differential proteins between these two groups, yielding 318 proteins which were differentially expressed between these groups (Sup Table 2). Group 1 underexpressed proteins related to translation and ribosomal processes and overexpressed proteins related to metabolism, specially glycolysis, T lymphocytes, and adhesion. On the other hand, Group 2 underexpressed proteins related to metabolism, T lymphocytes, and adhesion processes and overexpressed proteins related to translation and ribosomes (Figure 2).

**Figure 2:**
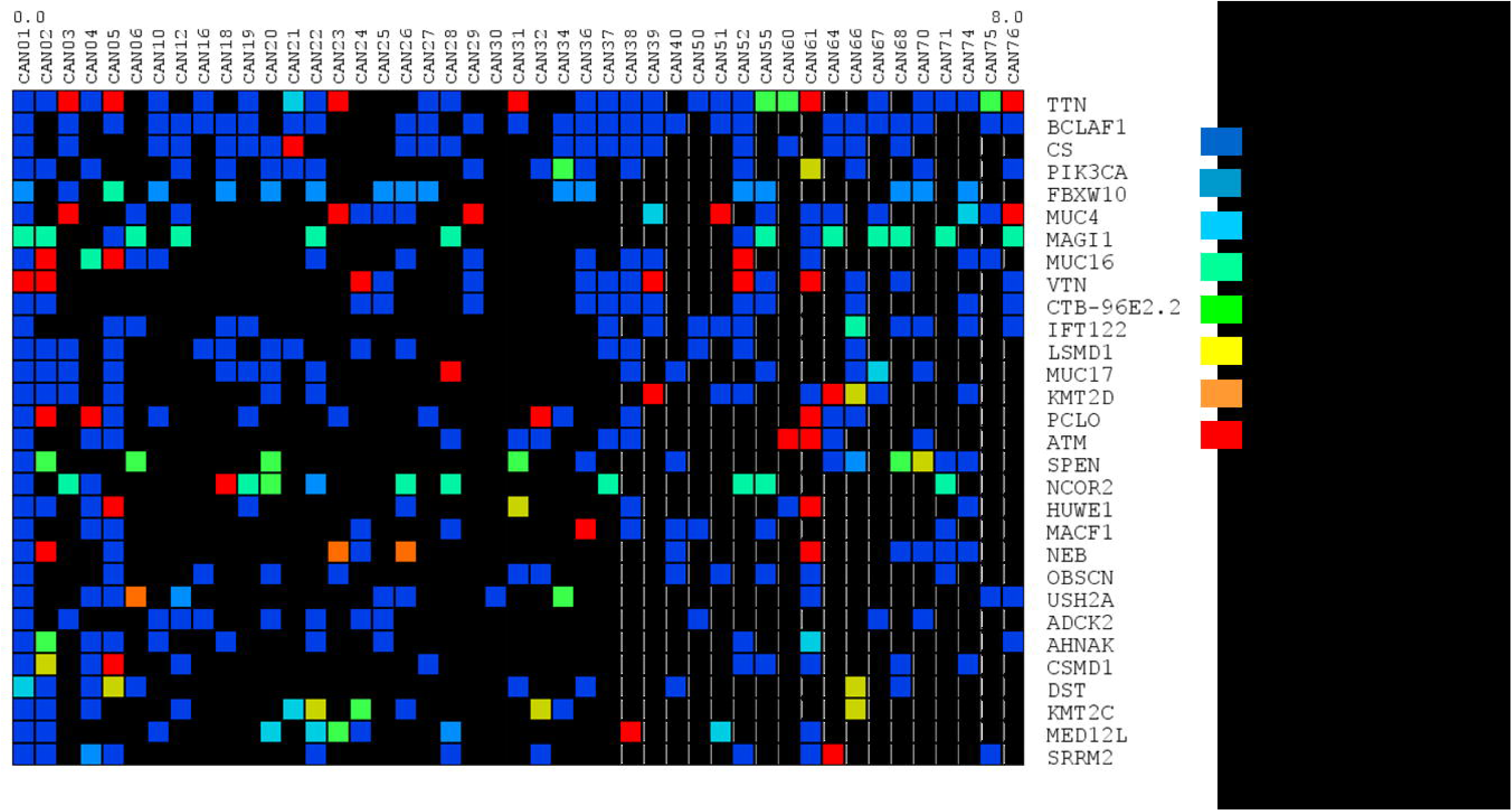
Significance Analysis of Microarrays identified 318 differential proteins between two groups of ASCC patients. Green=underexpressed. Red= overexpressed. In green, Group 1. In blue, Group 2.

With respect to the clinical data distribution between these two groups, both were comparable; there were no significant differences in the distribution of clinical parameters (Sup Table 3). In addition, there were not significant differences in disease-free survival or overall survival (Sup Fig 1).

A search in the Genomics of Drug Sensitivity in Cancer database (https://www.cancerrxgene.org/) suggested RAC1 (overexpressed in Group1 and underexpressed in Group 2) as a possible therapeutic target. The drug associated with this gene is EHT-1864.

### Functional characterization of proteomics data

To study proteomics data from a functional perspective, a PGM was created using the 1,954 proteins obtaining from the MS experiments. The resulting network was divided into 10 functional nodes, one of them with an overrepresentation of two biological functions (metabolism and mitochondria) (Figure 3).

**Figure 3:**
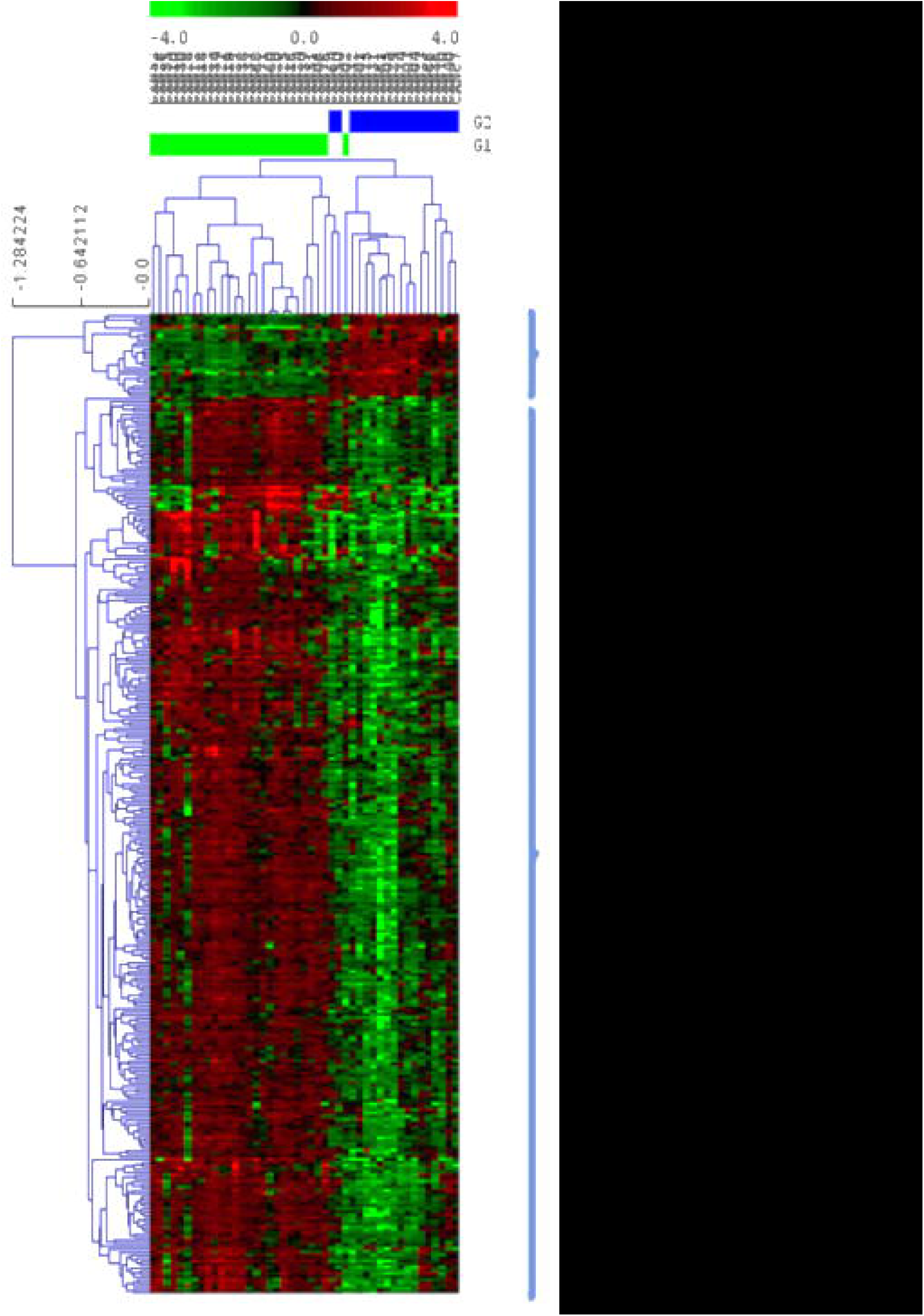
Functional network created using the proteomics data from the ASCC patients. Ten nodes with different biological functions were identified.

Then, functional node activities were used, as in previous works (7-9), to study the differences in biological processes between the two identified groups of patients. There were significant differences between the two groups in membrane category, in the two nodes related to metabolism, and in the nodes associated with adhesion, ribosomes, translation, extracellular matrix and splicing (Figure 4).

**Figure 4:**
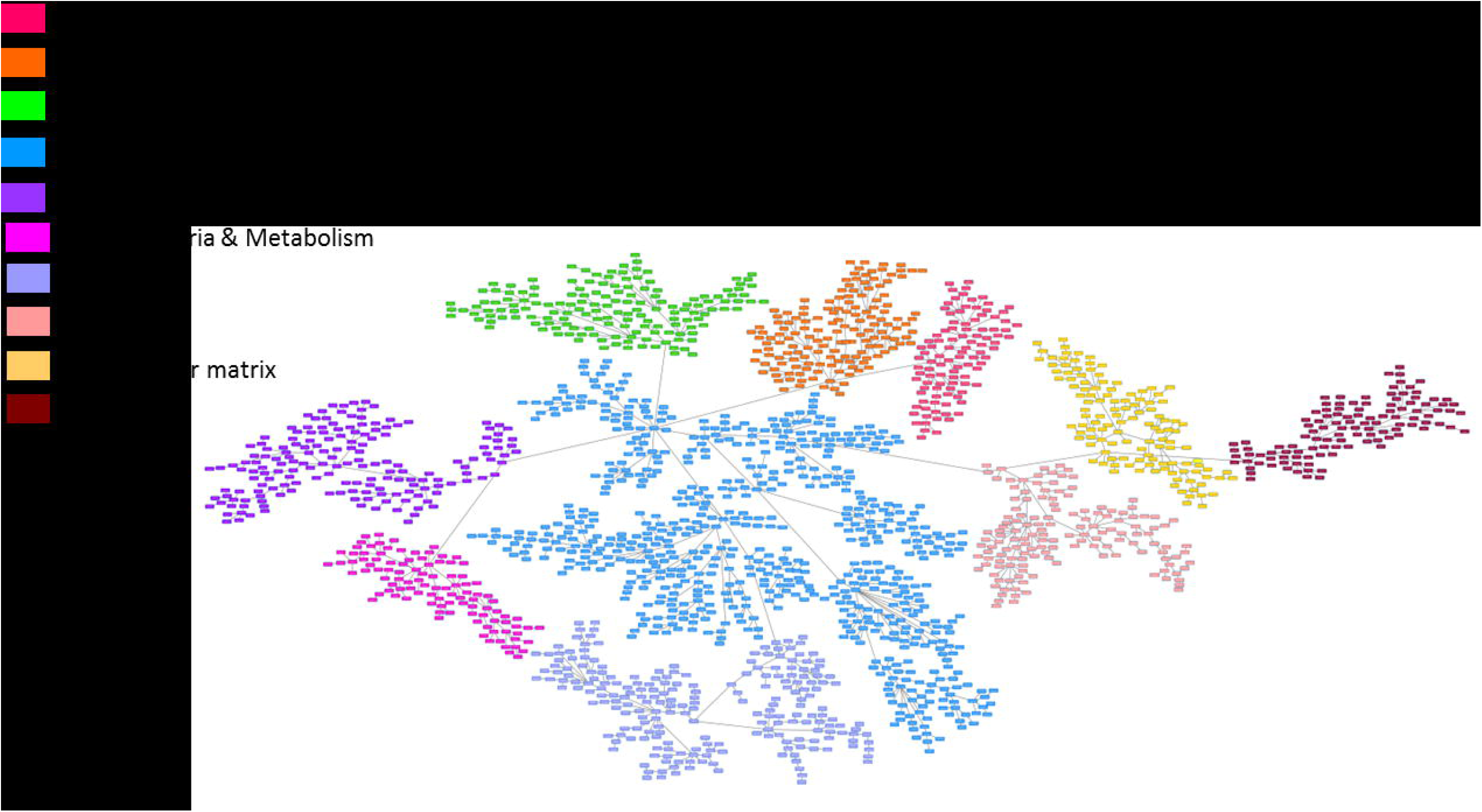
Functional node activities between the two groups defined by the hierarchical cluster. ****: p<0.0001; ***: 0.0001 ≥p≤ 0.001; **: 0.01≥p≤0.05. G1= Group 1; G2= Group 2.

### Metabolism nodes

We found two different nodes related to metabolism, both of which showed a higher expression in Group 1. The metabolism 1 node was formed by 158 proteins, mostly related to mitochondrial metabolism, especially the tricarboxylic acid cycle. Among them, three were also identified as differentially expressed by the SAM analysis: P09622 (DLD), P06744 (GPI) and P14550 (AKR1A1). The metabolism 2 node included 104 proteins related to mitochondria and metabolism, especially oxidative phosphorylation, such as P04406 (GAPDH), P06733 (ENO1), P07954 (FH) or Q9UI09 (NDUFA12).

### Adhesion node

The adhesion node showed a higher expression in Group 1. The adhesion node included 654 proteins, 25 of them classified by SAM as differentially expressed between the two groups of patients.

### Genetic variants with different frequencies between the two groups of patients established by proteomics

Using a Chi-squared test, the frequencies of genetic variants for each gene were compared to determine whether the groups of patients defined using proteomics data also showed differences in genetic variants. Twelve genes presented different frequencies of genetic variants between the two groups (Figure 5).

**Figure 5:**
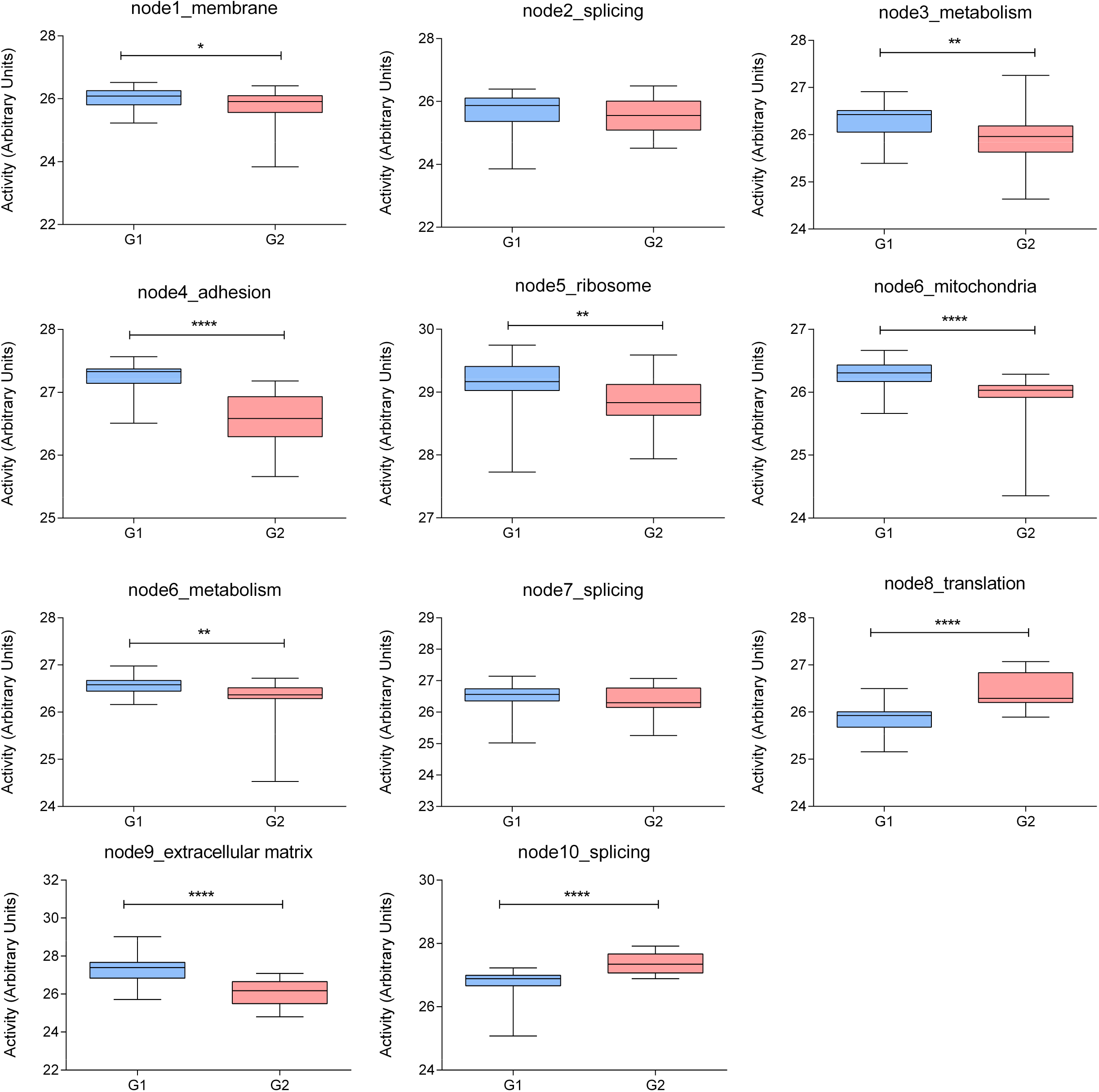
Genes with different frequency of genetic variants between the two groups of patients identified in proteomics. The mutation rate was calculated as the total number of mutations of a gene in each group divided by the number of patients assigned to that group.

### Tumor growth rate predicted by metabolic modeling

FBA allows for the comparison of the tumor growth rate between groups of tumors. The tumor growth rate predicted for Group 1 was significantly higher than the tumor growth rate predicted for Group 2 (Figure 6).

## Discussion

ASCC is an infrequent tumor. With no targeted therapy yet established, the molecular characterization of these tumors is still necessary. In this study, we combined the two main – omics, WES and proteomics, to further characterize a cohort of 46 patients diagnosed with primary ASCC. To our knowledge, this is the first study combining WES and proteomics in ASCC. The results of this study allow us to establish two molecular subgroups in ASCC with different molecular features. Moreover, the analyses of these two groups pointed out some drug-susceptible processes, such as metabolism, and suggested other possible therapeutic targets, like *ATM* and its relationship with PARP inhibitors (PARPi).

Previous studies have analyzed both primary and metastatic ASCC paraffin samples using WES or gene panels (10-13). A previous study using proteomics data to characterize different locations of anal cancer was also existed (14). However, this is the first study that performs proteomics experiments in localized ASCC samples and combines proteomics data with WES information. Previous WES studies served to identify frequently-occurring mutations in this disease, including mutations in the *PIK3CA, FBXW7, FAT1* and *ATM* genes (11-13). In our cohort, *PIK3CA* presented a genetic variant with a high or modifier impact in 40% of the patients, *FBXW7* in 16%, *FAT1* in 18%, and *ATM* in 27% of the patients.

Using proteomics data and HCL, it was possible to establish two different molecular groups of patients. Differential proteins were mainly related to the metabolism of glucose, translation and ribosomes, T lymphocytes and adhesion. Although these molecular groups have not been associated with any clinical or prognostic features, these processes may be relevant in the development of new therapeutic strategies. For instance, those tumors that overexpressed proteins related to glycolysis may be candidates for drugs targeting metabolism such as metformin, which has been shown to have cytostatic effects on other tumors, such as breast or bladder carcinoma (7). On the other hand, one of the main differences between these two groups is that Group 1 had a higher expression of proteins related to T lymphocytes. With the bloom of immunotherapies, immune proteins have acquired great relevance. Therefore, this group of patients may be good candidates for immunotherapy. In fact, nivolumab has been reported to be an effective therapy in metastatic ASCC and its efficacy is related to the presence of cytotoxic T cells (34). Moreover, pembrolizumab has demonstrated its antitumor activity in PD-L1-positive advanced ASCC (35).On the other hand, the search in Genomics of Drug Sensitivity in Cancer database to establish possible therapeutic targets suggested RAC1 (overexpressed in Group 1 and underexpressed in Group 2) as a potential therapeutic target. RAC1 has as associated drug, EHT-1864, which affects the cytoskeleton (36).

In addition, MS experiments and PGMs allow for the functional characterization of these two groups of patients. In this functional analysis, differences in metabolism were confirmed. There were also differences at the mitochondria level, which is the target for metformin. Metabolism nodes showed a higher expression in Group 1. P06744 (GPI, glucose 6-phosphate isomerase), included in the metabolism 1 node, is the enzyme that converts glucose – phosphate into fructose 6-phosphate and a higher expression has been associated with tumorigenesis and poor prognosis in gastric cancer (37).

The adhesion node also had a higher expression in Group 1 and contained 25 proteins identified by SAM as differentially expressed. P46940 (IQGAP1) has been associated with poor prognosis in head and neck squamous cell carcinoma (38) and has also been associated with response to chemo-radiotherapy in rectal adenocarcinomas (39). P27797 (CALR) induces an immune response in esophageal squamous cell carcinoma (40). P08133 (ANXA6) promotes EGFR deactivation (41). P62829 (RPL23) negatively regulates apoptosis and also inhibits growth in colorectal cancer (42). On the other hand, RPL23 has been identified as an oncogene in head and neck squamous cell carcinoma (43). O43707 (ACTN4) increases cell motility and invasion in colorectal cancer (44). Previous studies have described how P84077 (ARF1) forms a complex with EGFR and promotes invasion in head and neck squamous cell carcinoma (45). P35579 (MYH9) plays an important role in adhesion and migration, and its overexpression is correlated with metastasis in colorectal cancer through the MAPK pathway (46). Aberrant activity of P63000 (RAC1), which is involved in metastasis and proliferation, is a hallmark in cancer, (47). At the same time, O75131 (CPNE3) promotes cell migration through RAC1 (48). Finally, P61586 (RHOA), a tumor suppressor gene, plays a relevant role in colorectal cancer, being associated with metastasis and is deactivated in a significant number of colorectal tumors (49).In conclusion, the majority of the proteins included in this adhesion node play well-established roles in metastasis processes.

Moreover, the combination of the proteomics and genetic variants information showed that the two molecular groups defined by proteomics also had a different mutational profile. Group 2 showed a higher frequency of *ATM* genetic variants. Previous studies have described a high response rate to PARPi, as olaparib, in prostate tumors with mutations in *ATM* (50). Therefore, Group 2 patients may also be candidates for the treatment with PARPi.

In addition, FBA predicted a higher tumor growth rate for Group 1 than for Group 2. It may be possible that, given their higher proliferation, the tumors of Group 1 may also be more responsive to chemotherapy.

This study has some limitations. The results need to be validated in an independent cohort. The information in ASCC is scarce so a prospective validation will be needed. The number of proteins detected by MS still needs technical improvement to be considered to be at the same level as genomics. However, proteomics offers a more direct measurement of the effectors of biological processes. Finally, a consensus analysis pipeline to apply in cancer sequencing data is still necessary.

## Conclusions

In conclusion, two different molecular groups of patients have been proposed based on proteomics expression. This may be the first step toward a personalized therapy approach in ASCC. In addition, some possible targeted therapies, such as PARPi or immunotherapy, according to the molecular features (genetic and protein-based) defined in the two proteomics groups were suggested.

## Data Availability

The proteomics data generated during the current study are available in Chorus repository (https://chorusproject.org/pages/index.html). The exome sequencing data are available in SRA https://www.ncbi.nlm.nih.gov/sra) under the name PRJNA573670.

https://www.ncbi.nlm.nih.gov/sra

## List of abbreviations

ASCC: Anal squamous cell carcinoma
5FU: 5-fluorouracil
DFS: disease-free survival
WES: whole-exome sequencing
MS: mass-spectrometry
PGM: probabilistic graphical model
FFPE: formalin-fixed paraffin-embedded
ECOG-PS: Eastern Cooperative Oncology Group performance status
HPV: Human papilloma virus
VEP: Variant Effect Predictor
DDA: Data-dependent acquisition
FDR: False Discovery Rate
FBA: Flux Balance Analysis
BIC: Bayesian Information Criterion
GPR: Gene-Protein-Reaction rules
SAM: Significance Analysis of Microarrays
PARPi: PARP inhibitors

## DECLARATIONS

### Ethics approval and consent to participate

This study was approved by the Ethics Committee of Hospital Universitario La Paz (PI-1926). Informed consents were obtained for all patients included in this study.

### Consent for publication

Not applicable.

### Authors’ contributions

All the authors have directly participated in the preparation of this manuscript and have approved the final version submitted and declare no ethical conflicts of interest. RL-V, MM, EL-C, and VH contributed the DNA and protein extraction. MM and VH contributed the HPV determination. LG-P, CP, and MC contributed the pathological review of the samples. PN and CF contributed the mass spectrometry data. RR-R and CL contributed the sequencing analyses. LT-F, AG-P and JAFV contributed the probabilistic graphical models. LT-F contributed the FBA analyses. IG, JM, PG-A, JC, CC, RG-C, and JF contributed the clinical data and the analyses related. LT-F, IG, AG-P, GP-V, and AZ-M contributed in the design of the study and the statistical and gene ontology analyses. L-T-F drafted the manuscript. IG, JM, JAFV, and JF conceived of the study, and participated in its design and interpretation. AG-P, JM, JAFV, and JF reviewed the manuscript. JF coordinated the study. All authors read and approved the final manuscript.

## TABLE AND FIGURE LEGENDS

Figure 6: Tumor growth rate predicted using metabolic models. a.u. = arbitrary units. *: p <0.05

Sup Fig 1: A. Disease-free survival curves for the two groups defined by proteomics. B. Overall survival curves for the two groups defined by proteomics.

Table 1: Patient characteristics.

Sup Table 1: Genes with a detected genetic variant in at least 10% of the patients.

Sup Table 2: 318 differential proteins obtained by SAM between the two groups.

Sup Table 3: Clinical data distribution between the two molecular groups. p= p-values obtained from a Chi-squared test comparing the two groups of patients.

